# Association between Neonatal Necrotizing Enterocolitis and Neurodevelopmental Outcomes: A Comprehensive Systematic Review and Meta-Analysis

**DOI:** 10.1101/2024.09.20.24314095

**Authors:** Jonathan A. Berken, Priyanka Ramanan, Mary J. Akel, Corinne Miller, Denise Nunes, Daniel N. Aleynick, Joshua B. Wechsler, Lauren S. Wakschlag, Leena B. Mithal

## Abstract

Preterm infants are at high risk for systemic inflammatory disorders, including sepsis, meningitis, bronchopulmonary dysplasia, and necrotizing enterocolitis (NEC). The developing brain of the premature newborn is especially susceptible to the cascade of inflammatory mediators elaborated in these conditions that cross the blood-brain barrier. NEC, a severe and potentially fatal condition of the gut that occurs in premature newborns, is a prime example of how an inflammatory reaction, perhaps initially localized, can become generalized and cause systemic harm. One such result is brain injury, especially to the cerebral white matter, which may lead to neurodevelopmental abnormality and dysregulated behavior. Numerous studies have documented an association between necrotizing enterocolitis and neurodevelopmental impairment (NDI), but to date, the brain and behavioral deficits associated with neonatal NEC are not fully understood. We performed a comprehensive systematic review and meta-analysis of existing literature to characterize brain injury and behavioral alterations associated with NEC. 7153 peer-reviewed published manuscripts were screened by two independent reviewers and evidence quality was assessed using GRADE criteria. Of these papers, 62 satisfied the criteria for our review (i.e., no case reports, meta-analyses, systematic reviews, or animal studies). Data from 32 papers using Bayley Scales of Infant and Toddler Development to assess infant outcomes were included in the meta-analysis. Our findings support neonatal necrotizing enterocolitis having deleterious effects on brain and behavioral development and impact on cognitive function, risk of cerebral palsy and motor impairment, educational achievement, behavior, and neuroanatomy. We discuss herein findings of both short-term outcomes (1-3 years) and long-term outcomes (until 13 years). Our meta-analysis also indicates that NEC has a moderate effect size on infant development, with consistent impairment across mental, cognitive, language and motor domains.

**WHAT DO THE FINDINGS OF THIS REVIEW MEAN?:** In this review, we describe several short-term and long-term neurodevelopmental outcomes of preterm infants with NEC. These findings suggest that infants with NEC should be monitored with close developmental follow-up so that individuals can receive appropriate testing that may prompt therapies and qualify them to receive early intervention services.

**HOW UP TO DATE IS THIS REVIEW?:** The review authors searched for studies published up to 2021. Abstracts with no data available were excluded, so this review does not consider any new findings published from 2022-2024.

## BACKGROUND

Necrotizing enterocolitis (NEC) is a severe inflammatory disease of the small intestine and colon that affects between 3% to 15% of very low (< 1,500 g, VLBW) and extremely low (< 1,000 g, ELBW) birthweight preterm neonates.^1^ The disorder is characterized by inflammatory changes in the gut, ranging from mild mucosal injury to tissue necrosis and perforation. NEC is stratified into three groups by the modified Bell staging criteria, based on clinical presentation (**Table 1**).^1^ Such classification allows prognostication about disease sequelae and is critical to therapeutic decision making. Stage I refers to mild or suspected NEC whereas Stage II-III have proven abnormalities radiographically and with increasing severity of illness. Stage IIIb NEC necessitates surgical intervention and has a mortality rate of 50% despite treatment.^2-5^ Evidence suggests a multifactorial pathogenesis of NEC with contributing factors of intestinal immaturity, gut dysbiosis, infection, and formula feeding.^6-13^

**Table 1.**
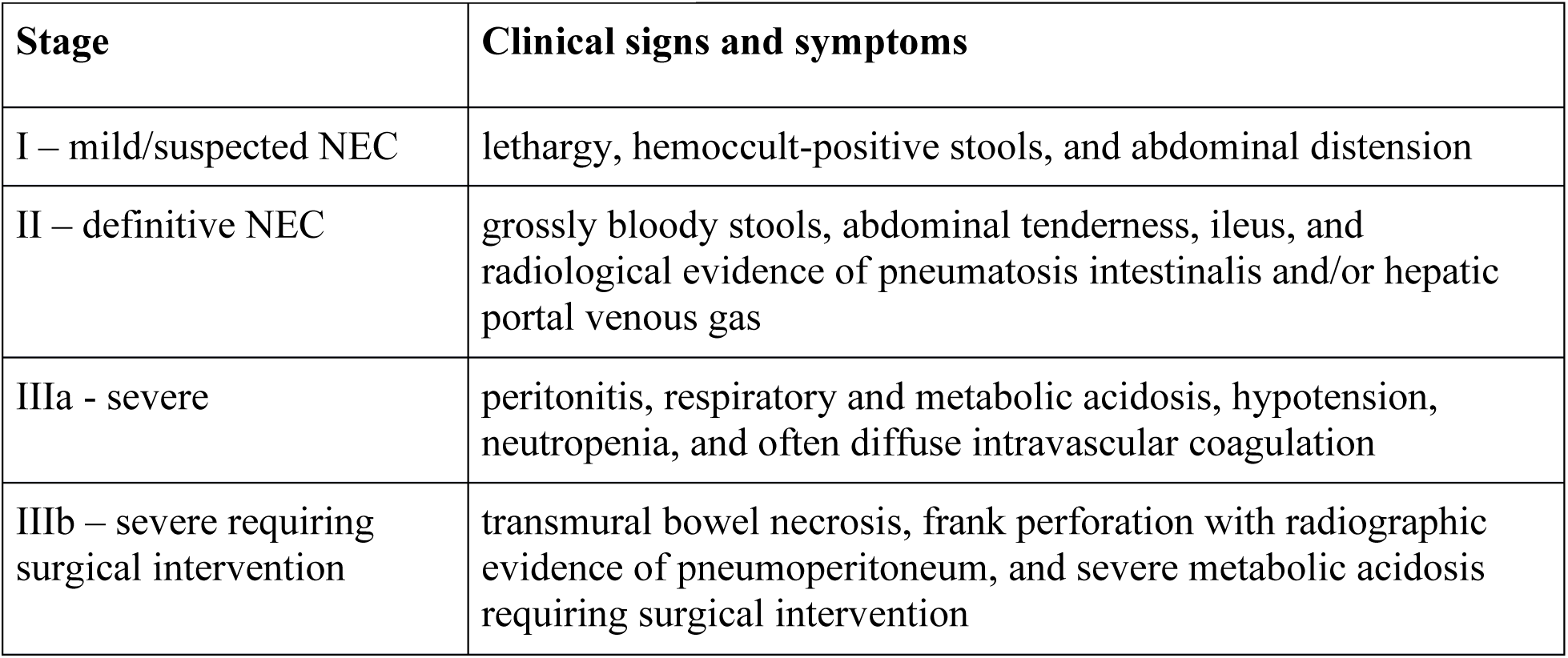
Bell staging criteria.

| Stage | Clinical signs and symptoms |
| --- | --- |
| I – mild/suspected NEC | lethargy, hemocult-positive stools, and abdominal distension |
| II – definitive NEC | grossly bloody stools, abdominal tenderness, ileus, and radiological evidence of pneumatosis intestinalis and/or hepatic portal venous gas |
| IIIa - severe | peritonitis, respiratory and metabolic acidosis, hypotension, neutropenia, and often diffuse intravascular coagulation |
| IIIb – severe requiring surgical intervention | transmural bowel necrosis, frank perforation with radiographic evidence of pneumoperitoneum, and severe metabolic acidosis requiring surgical intervention |

While the disorder has its origin in the premature gut, NEC is a systemic disease and, thus, the developing neonatal brain is particularly vulnerable to harm.^6^ Studies point to potential inflammatory triggers in NEC. For example, products of a dysbiotic gut flora, particularly lipopolysaccharides elaborated by an overabundance of *Proteobacteria* species, promote toll-like receptor 4 (TLR4) activation on the intestinal epithelium. TLR4 activation, in turn, induces the production of the transcription factor NFkβ and the release of injurious proinflammatory cytokines that promote intestinal infiltration by CD4+ T-cells, initiating gut injury.^14^ In addition, the proinflammatory molecules released as the result of TLR4 activation, which are found in the plasma of NEC infants as well as in intestinal tissue, traverse the porous blood-brain-barrier of the premature neonate along with CD4+ T-cells leading to microglial activation, the accumulation of damaging reactive oxygen species, neuronal loss, oligodendrocyte damage, and myelin deficit.^14,15^ While brain injury in infants with NEC primarily involves the white matter, manifesting most commonly as periventricular leukomalacia, reduced gray matter volume also occurs in the thalamus, basal ganglia, and cerebellar dentate nuclei.^16-19^ Neurogenesis, neuronal migration, scheduled apoptosis, and synaptogenesis occur in the second and third trimesters, and thus the preterm brain is particularly susceptible to inflammation at critical periods of neurodevelopment.

In view of the observed harmful effects of inflammatory mediators, both molecular and cellular, on the brains of neonates with Stage II and Stage III disease, it would be plausible that NEC survivors will show some degree of neurodevelopmental impairment (NDI) later in childhood. Several studies, in fact, have identified NEC as an independent risk factor for NDI in children born preterm, above and beyond the risk of brain trauma associated with premature birth itself.^14^ In addition, there are data that infants requiring surgery for NEC fare more poorly neurodevelopmentally than those treated medically, a comparison that appears to remain significant when compared to infants undergoing abdominal surgery for other conditions.^20-22^ Not all studies, however, support these observations, as differences in degree of prematurity, heterogeneity of NEC severity among subjects, age of behavioral testing, testing inventories used, and the number of infants assessed make definitive inferences about the relationship between NEC and NDI difficult.

Here, we apply a holistic framework to systematic review and meta-analysis to determine how NEC alters development and behavior across multiple functional domains such as motor skills, cognition and language, IQ, school achievement, and social-emotional performance. What sets this paper apart is its methodology. We comprehensively screened thousands of manuscripts, including only high-quality peer-reviewed evidence-based studies, to describe neurodevelopmental outcomes of NEC that extend beyond traditional measures. Further, this study seeks to assess how factors such as NEC severity, gestational age, and age at testing influence the degree of NDI and to establish whether NDI persists into later stages of childhood. These results and ultimate conclusions can pave the way for targeted interventions and improved clinical care for survivors of NEC.

## OBJECTIVES

The objectives of this study were to 1) describe and summarize the impact of NEC on neurodevelopment outcomes including: cognitive function, motor impairment, educational achievement, neuroanatomy, social-emotional behavior, and long-term behavioral and language outcomes, and 2) perform a meta-analysis to determine the strength of association between NEC and neurodevelopmental impairment.

## METHODS

### CRITERIA FOR CONSIDERING STUDIES FOR THIS REVIEW

The current systematic review and meta-analysis were conducted as part of a larger study exploring the relationship between systemic inflammation in prematurely born infants and their neurodevelopmental outcomes. This broader investigation aimed to understand how various inflammatory conditions could influence the developing brain in this vulnerable population. However, to provide more specific insights, the focus was narrowed to necrotizing enterocolitis. NEC was chosen because it is a significant and severe inflammatory condition unique to premature infants, with impact on neurodevelopment. Despite this, the specifics of how NEC affects brain and behavioral maturation are not clearly defined, as premature populations are often very heterogeneous, and the care provided within different neonatal intensive care units (NICUs) varies. Therefore, we aimed to take a comprehensive approach, acknowledging the diversity within this population and the complexities of their care.

### SEARCH METHODS FOR IDENTIFICATION OF STUDIES

Two medical librarians (CHM, DN) searched Embase (Elsevier), Cochrane Library (Wiley), CINAHL (EBSCO), PsycINFO (EBSCO), and PubMed (NIH/NLM) from inception date through the date of the search, January 20, 2022. Keywords and subject headings were used to search for literature on necrotizing enterocolitis or systemic inflammation and neurodevelopment in newborns. The full search strategies are available [Supplementary Material]. After collation and de-duplication in Endnote, records were uploaded to Rayyan for title and abstract screening.

### SELECTION OF STUDIES

Articles were uploaded to Rayyan, a web-based tool for systematic reviews, for an initial screening. Two independent reviewers JB and PR meticulously reviewed all the titles and abstracts to identify those relevant to the broader study focusing on systemic inflammation and brain development in prematurely born infants. During this first review, articles that specifically addressed systemic inflammation and its impact on brain development were included, while duplicates were removed. Additionally, case studies, case series, and reviews were excluded from further consideration (see below for further details on exclusion criteria). Any disagreements between JB and PR regarding the inclusion of specific articles were discussed and resolved in a follow-up session. For any ongoing disagreements, a final decision was made by LM, ensuring consistency and accuracy in the selection process.

In the current study, we further categorized the selected papers based on whether they contained data specifically linking necrotizing enterocolitis to developmental outcomes. This focused categorization was crucial in refining our understanding of how NEC, as a distinct inflammatory condition, might influence neurodevelopmental trajectories in premature infants. From the initial pool of articles, our title and abstract review identified 140 papers that met these specific criteria, highlighting the connection between NEC and developmental outcomes. This targeted selection allowed us to concentrate on the most relevant literature, providing a clearer picture of NEC’s unique impact on neurodevelopment.

### TITLE AND ABSTRACT REVIEW

The articles identified from all electronic databases were evaluated for duplicates to ensure each was a unique manuscript. Next, title and abstract review was conducted by two team members (JB, PR) to evaluate for inclusion. Papers not clearly excluded through title and abstract review underwent article extraction review.

### INTER-RATER RELIABILITY

Ten initial papers were reviewed by the study team (JB, PR, MA) to ensure consensus and validate the extraction form. After discussion and identification of discrepancies, the standardized extraction form was edited, and three additional articles were reviewed by the study team to ensure reliability.

### FULL TEXT EXTRACTION

Each included article was reviewed twice by two different study team members (JB, PR, MA). For any articles where the first and second reviewers disagreed, a third reviewer completed data extraction to resolve the discrepancy. Data from the articles were extracted into a standardized REDCap form developed by the study team. The standardized form prompted extraction of the following elements: study characteristics (design, study population, timepoints, etc.), outcome measures, and results.

### DATA ANALYSIS AND SYNTHESIS

#### SYSTEMATIC REVIEW ANALYSIS AND INVESTIGATION OF HETEROGENEITY

Articles were qualitatively evaluated to determine heterogeneity across key extraction components, including sample demographics, outcome measures, and results. Following qualitative evaluation, data were quantified and summarized across these same components. Given the heterogeneity of the data, articles were grouped by outcome measure for review analysis. Data reporting was conducted per PRISMA checklist.

### META-ANALYSIS METHODS

Each article was assessed by statisticians (AM, SA) on the following criteria to be included in the meta-analysis: 1) utilized Bayley Scales of Infant and Toddler Development (Third or Second Edition), 2) directly compared those with NEC to those without NEC, 3) only focused on infant development as the outcome. Following this, a Cohen’s d value for effect size and subsequent standard error was calculated for each individual study to assess the magnitude of the effect of NEC on infant development. Effect size was calculated via a comparison of two proportions or mean difference that is detailed in Cohen.^23^ When proportions or means were not directly reported in the studies included, Cohen’s d values were derived using alternative statistics, specifically odds ratios or unstandardized regression coefficients. Cohen’s d values were then estimated from the odds ratios or unstandardized regression coefficients using an online calculator or computational tool.^24^

We utilized a random-effects model for the meta-analysis to account for potential heterogeneity among the included studies. The random-effects model assumes that the true effect size may vary across studies and incorporates both within-study and between-study variability. Our primary outcome of interest was the standardized mean difference (SMD) between those with NEC and those without, calculated by pooling the individual effect sizes weighted by the inverse of the variances. Heterogeneity was assessed among studies utilizing Cochran’s Q test and the I2 statistic. Significance was set at p < 0.05 for Cochran’s Q test. All statistical analyses were conducted using R version 4.1.2. Forest plots were generated utilizing the meta package in R.^25^

### ASSESSMENT OF QUALITY AND RISK OF BIAS

Quality and risk of bias in the final set of articles was assessed by applying the Grading of Recommendations Assessment, Development and Evaluation (GRADE) framework. GRADE is a systematic approach to rating the certainty of evidence in systematic reviews and other evidence syntheses.^26^ To achieve the goal of rating the quality of evidence for each outcome presented in a study, and to also provide a recommendation of how to dispose of that evidence, GRADE uses a category system to assign ratings to evidence ranging from “very low” to “high”.^27^ GRADE also uses a five-domain system for rating the quality of evidence, including: 1) Risk of Bias, 2) Inconsistency, 3) Indirectness, 4) Imprecision, and 5) Publication Bias.^27^ At least two study team members assessed each article using the GRADE framework; for any articles where there was discordance in the first two reviewers’ GRADE scores, a third reviewer assessed to resolve the disagreement. Only those articles that received a GRADE score of “moderate” or higher were included in the final analyses.

## RESULTS

### DESCRIPTION OF STUDIES INCLUDED IN REVIEW AND RESULTS OF SEARCH

The preliminary database search initially identified 9,644 manuscripts, which were reduced to 7,153 peer-reviewed papers after removing duplicates (**Figure 1**). Following a screening process for relevance to NEC and neurodevelopment, and a thorough quality assessment, this number was narrowed down to 67 articles. Of these, 37 manuscripts were included in the meta-analysis. These studies report on cognitive, motor, neuroimaging, and educational outcomes, with follow-up periods ranging from 18 months to 30 years.

**Figure 1.**
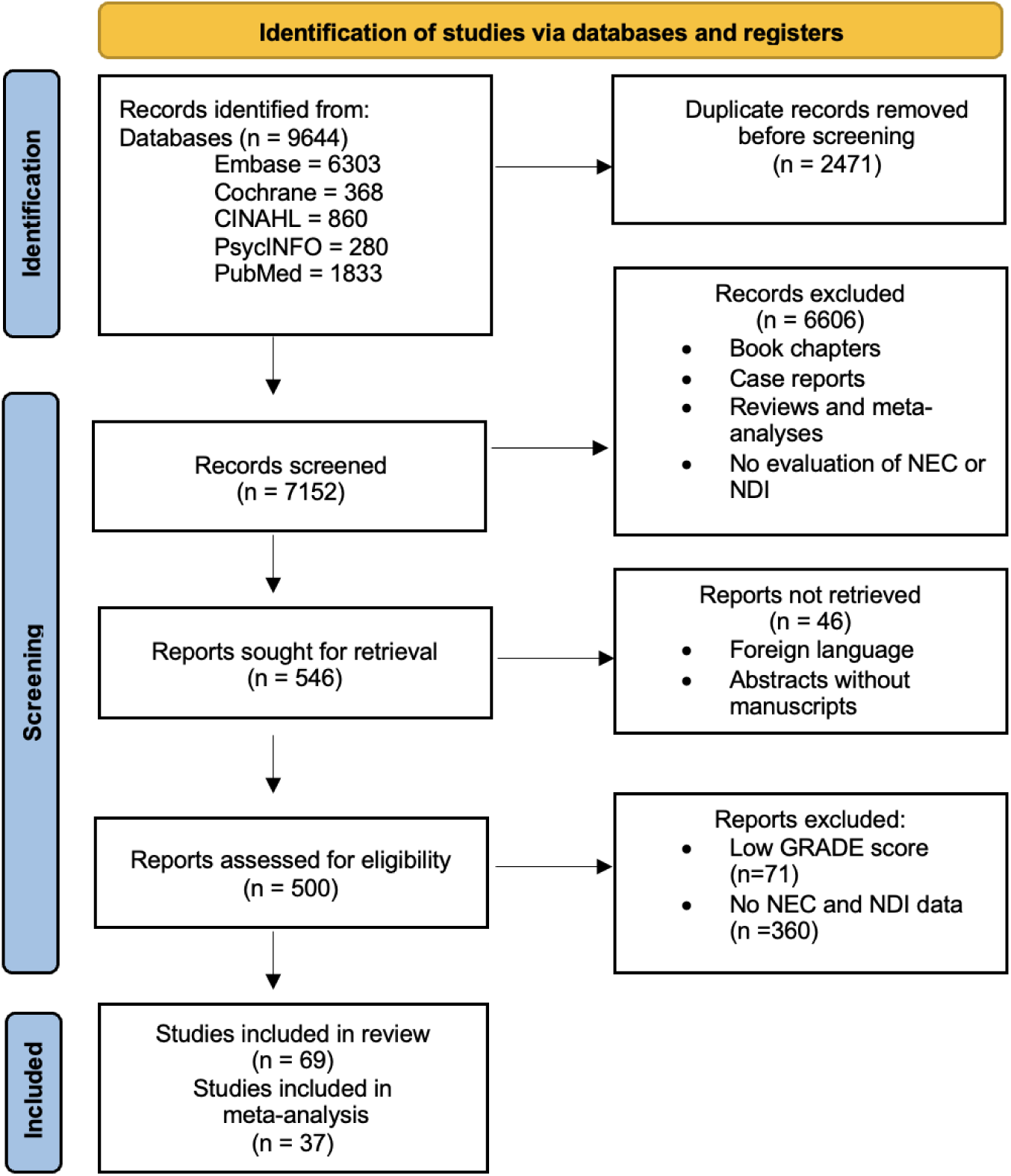
PRISMA identification of studies for inclusion. This figure presents the study selection process for the investigation of associations between neonatal necrotizing enterocolitis in premature infants and neurodevelopmental outcomes. Initially, studies were identified through comprehensive searches of databases and registers. Following identification, records underwent a rigorous screening process to assess eligibility. Quality of the studies was evaluated using the GRADE scale. Finally, studies meeting the eligibility and quality criteria were included in both the systematic review and meta-analysis.

### SYNTHESIS OF RESULTS

#### MOTOR OUTCOMES

##### Cerebral palsy

More evidence suggested no significant increase in the rates of cerebral palsy among survivors of NEC compared to infants with no history of NEC. The earliest study included, reporting CP rates in 1995, involved a relatively small cohort (n=89) and found no differences in rates of cerebral palsy, as assessed at three and five years, between infants with NEC requiring surgery and age-matched controls.^28^ The absence of an association seemed corroborated by subsequent larger cohort studies. Notably, Pike et al., 2012, in a large cohort study (n=6618) among ELBW infants who developed cerebral palsy, found the odds of having NEC to be non-significant (OR= 1.23, [0.58-2.63]).^29^ More recently, Humberg et al., 2020, in another large cohort study (n=2241) of VLBW infants in the German neonatal network between 22-36 weeks’ gestation, found no significant rates of CP (p=0.289).^30^

This conflicted with other data, notably Martin et al., 2010, suggesting that the risk of CP was higher among infants with co-existing NEC and bacteremia.^31^ This study found that children with late bacteremia and surgical NEC were ∼eight times more likely to develop diparetic cerebral palsy, while children with late bacteremia and medical NEC were twice as likely to develop diparetic cerebral palsy, compared to children with neither NEC nor bacteremia.^31^ This was supported by Thome et al., 2017 (n=233), who found a significant association between a higher GMFCS score and NEC,^32^ and by Waugh et al., 1996 (n=199), who found the odds of NEC in infants with CP to be 3.0 [1.0-9.1] on univariate analysis.^33^ This association became less significant in multivariate analysis, although sample size might potentially explain this difference.

##### Psychomotor development (Bayley Assessment)

There was overwhelming evidence that preterm infants with NEC had significantly lower psychomotor scores as assessed through BSID-II or III compared with age-matched controls without a history of NEC and controlling for co-morbidities. Larger cohort studies of infants <30 weeks GA found that surgical NEC had increased odds of NDI, including lower Psychomotor Developmental Index (PDI) components of Bayley (p <0.0001).^34^ Surgically treated NEC showed significantly lower PDI scores compared with conservative treatment across multiple studies.^31,35-37^ A diagnosis of NEC, regardless of treatment, also seemed to confer a risk of lower PDI scores.^32,38-40^ Yeh et al., 2004 also found that the lower PDI scores among infants with NEC found at 6 months of age persisted at 18 months as well (while Mental Developmental Index [MDI] scores improved with time).^41^ Additionally, a longer time to full-enteral feeding post-NEC was also associated with significantly lower Bayley motor scores, after adjusting for GA, LOS, and surgical intervention.^42^ However, there were two studies that did not find an association between abnormal PDI scores and NEC.^43,44^

##### Motor impairment from other assessments

There was less evidence to support a relationship between NEC and motor impairment as assessed using non-Bayley measures, although the diversity in motor scales used for assessment inherently complicated comparisons across studies. There seemed to be weaker focal motor differences among infants with NEC across various studies. Brown et al., 2006, using the Hammersmith Neonatal Neurological Assessment, found that when comparing <30 weeks GA infants to term infants, NEC was associated with altered tone (p=0.008) and abnormal tone patterns (p=0.020).^45^ Muto et al., 2021 found that NEC had an overall downtrend in postural-motor development scores, as assessed using the Kyoto Scale of Development, although the number of infants with NEC in this study was small (n=9).^46^ Shin SH et al., 2021 found abnormal gross motor (p=0.012) and fine motor scores (p=0.043) when comparing infants with NEC vs SIP using the K-ASQ; but these differences did not hold when assessed using the BSID-II.^47^ Sonntag et al., 2000 found no significant differences in psychomotor retardation.^48^ This was supported by Taylor et al., 2006, who utilized the Test of Motor Proficiency,^49^ and Salavati et al., 2021, who utilized Prechtl’s method of assessing early motor repertoire.^50^ There was no significant difference in motor skills when investigating infants with NEC who underwent enterostomy compared to primary anastomosis.^51^

#### COGNITIVE OUTCOMES

Of the 39 full-text published articles on necrotizing enterocolitis and neurodevelopment that satisfied the GRADE requirements, 16 reported data that specifically assessed the association between NEC and cognition. Of these, 12 (75%) found significantly lower scores achieved by NEC survivors versus age-matched controls without NEC on tests of cognitive performance.^18,31,36,41,46,47,52-54^ Four studies found no difference between these groups in this neurodevelopmental domain.^37,42,55,56^ Of these, three had relatively small subject pools (Ntotal < 50),^42,55,56^ including one paper with only 18 total subjects.^55^ Twelve of the papers reported data relating to NEC and cognition based on the MDI sub-scores on the Bayley Scales of Infant and Toddler Development (BSID II or III), evaluated in follow-up as early as 12 months to as late as 36 months.^19,32,34,36,38,43,53,56-60^ One study looked at cognitive performance in infants who had undergone surgery for NEC (Stage IIIb) and correlated the sequela of the associated white matter brain injury (WMBI), as detected by MRI, with MDI scores on BSID III.^18^ Four studies found significantly lower cognitive scores in NEC infants compared to controls using other well-validated inventories, including the Kyoto Scale of Psychological Development^46^ and the Gesell Development Schedules.^47^

#### BEHAVIORAL OUTCOMES

Eight of 69 studies included at least one behavioral outcome measure.^29,42,48,61-65^ These measures included attention disorders,^29,61,62,65^ assessment of inhibition,^63^ scores from the Child Behaviour Checklist,^29,42,66^ scores from the parent-reported Strengths and Difficulties Questionnaire,^29,64,67^ and social behavior.^48,65^ Results from studies with behavioral outcomes were somewhat mixed. Four articles found a significant association between NEC and behavioral outcomes.^48,61,63,65^ Astbury et al, showed that children with ADD at two years of age were significantly more likely to have NEC (p<0.0001) compared to those without ADD.^61^ Sonntag et al., found that among infants with NEC, subscales that of the Griffiths III that assessed personal and social skills were significantly lower (p=0.003) than those without NEC at 20 months corrected age.^48^ Differences in behavior outcomes were seen both in younger children (24 months or less) as well as school-aged kids. Taylor et al., studied behavioral outcomes among elementary school aged children at who were around seven years of age; they found significant increases in scores of parent-reported hyperactivity among children who had NEC.^65^ Nyman et al., conducted assessments at 11 years of age and showed that children with NEC scored significantly higher in the Inhibition Index (i.e., there were more concerns for the child’s ability to resist impulses) compared to those without NEC (p=0.014).^63^ However, the other half of studies found no significant associations between NEC and adverse behavioral outcomes.^29,42,62,64^

##### Attention Deficit Disorder

More recent evidence suggested that there did not seem to be a significant relationship between NEC and an increased risk of ADHD.^29,62^ Earlier studies had reported NEC as a significant predictor of increased hyperactivity and lower adaptive behavior scores after adjusting for social risks and predicted cognitive ability based on their neonatal risk profile.^65^ Astbury et al., 1995 also reported that children with ADD tended to have significantly more NEC (p <0.0001).^61^

#### EDUCATIONAL IMPACT

The few studies that investigated neonatal NEC and later educational outcomes had contradicting results. Johnson et al., 2011 investigated outcomes up to 11 years of age and found NEC to be a significant independent predictor of educational deficits, accounting for a 24-point deficit in standardized scores in reading (p=0.016) and math (p=0.002).^68^ Taylor et al., 2006 investigated NEC impact on subset domains of academic skills, finding significant odds of NEC among students at mean age of six years, with deficits in skills such as calculation (p=0.003) and academic skills cluster (composite of reading, writing and math) (p=0.049) scores.^49^ These findings contradicted the ORACLE Children study,^29^ which found no significant relationship between neonatal NEC and educational outcomes at seven years (across reading, writing, and math scores). No significant differences were noted between neonatal NEC and scores from parent and teacher rating scales, specifically the Strengths and Difficulties Questionnaire.^64^

##### Language

With regards to language, Chen et al., 2011 specifically found that students with a history of NEC have significantly lower language subdomain scores measured by the Gisell Developmental schedules (68 [SD=12.65] vs. 95 [SD=11.41], P<0.001).^52^ Of note, the same study noted inferior scores across other domains including gross motor, fine motor and adaptive behavior. In a study of infants with surgical NEC, those with white matter brain injury (WMBI) had significantly lower language scores (58.4 ± 14.3 vs. 70.6 ± 6.4; p = 0.082) on neurodevelopment assessment compared to infants without WMBI.^18^ A study examining the postoperative outcomes and developmental index of ELBW infants with NEC between the ages 1.5 - 3 years found that none of the children (n=9) had reached the expected developmental score for social language domain, with scores of 68.4 at 1.5 years corrected age and 69.1 at 3 years - scores that are considered poor.^46^

##### Intelligence

Of the studies that investigated trends in intellectual impairment, there was a common trend of lower IQ scores among infants with NEC when tested at school age. Humberg et al., 2020, in their cohort study of the German neonatal network, found that infants with a history of NEC had a threefold increased risk (RR 3.0 [1.8-4.2], p < 0.001) of developing IQ scores <85, which continued to show significant differences in subsequent matched-cohort analysis.^30^ Interestingly, a history of surgical SIP was not associated with lower IQs at school age. Similarly, Waugh et al., 1996, reported an increased odds ratio of NEC in infants with intellectual impairment (5.3 [2.0 - 14.0]) as assessed using the Griffith General Quotient, although this effect was not significant on multivariate analysis.^33^ In a smaller study of siblings, Stevenson et al., 1980, found significantly lower scores in NEC survivors compared with matched siblings.^69^ Additionally, a smaller study by Ta et al., 2010, that investigated the differences in outcomes among infants with NEC who received an enterostomy found that they scored significantly worse on intelligence scales compared to children who received primary anastomosis.^51^

#### NEUROIMAGING AND NEUROANATOMY

##### NEC and MRI Findings

Three papers examined the effects of NEC on brain structural integrity as seen on MRI.^18,52,54^ One paper reported significant diminution in the volume of both the gray and white matter in the brains of NEC survivors compared with matched controls, most conspicuously in the white matter.^18^ The infants with NEC-associated WMBI had lower MDI scores than those without WMBI. In addition, a study using both MRI and EEG to compare the brain structure and function of NEC survivors and age-matched children without a history of NEC, reported a statistically greater prevalence of abnormalities in the NEC group that were associated with significantly lower scores on the Gesell Development Schedules in the cognitive domains of adaptive behavior and personal-social function.^52^ Similarly, an investigation of the white matter of NEC survivors found significant differences in microstructure when compared with controls, particularly in the splenium of the corpus callosum, that were associated with statistically lower MDI scores on the BSID III.^54^

#### META-ANALYSIS

A total of 16 unique studies had Bayley-II scores or a combination of both Bayley-II and Bayley-III scores, while a total of 7 unique studies had Bayley-III scores. For the Bayley-II score, the pooled effect size for the standardized mean difference between individuals with NEC and those without was calculated to be 0.44 (95% CI: 0.32 to 0.55) (**Figure 3**). This indicates a moderate effect size, where individuals with NEC have Bayley-II scores lower than those without NEC. The I^2^ statistic was calculated to be 71%, suggesting moderate heterogeneity among the studies included in the meta-analysis.^70^ Cochran’s Q test yielded a significant result (Q = 93.03, p < 0.001), indicating heterogeneity among the studies. The results for the other analyses, including Bayley-III (**Figure 4**), composite Bayley-II and Bayley-III (**Figure 2**), Bayley subscores (motor, mental, cognitive, and language) are presented in the **Table 2**, detailing the confidence intervals, effect sizes, I² statistics, and Cochran’s Q test with p-values.^70^ Data related to the composite Bayley-II and Bayley-III scores are also stratified based on whether the infants were born extremely preterm or very preterm and whether testing occurred before or after 18 months of age. The 18-month threshold was selected because it aligns with the typical schedule of follow-up clinics, which often conduct developmental assessments at this age to provide an early evaluation of cognitive and motor development.

**Figure 2.**
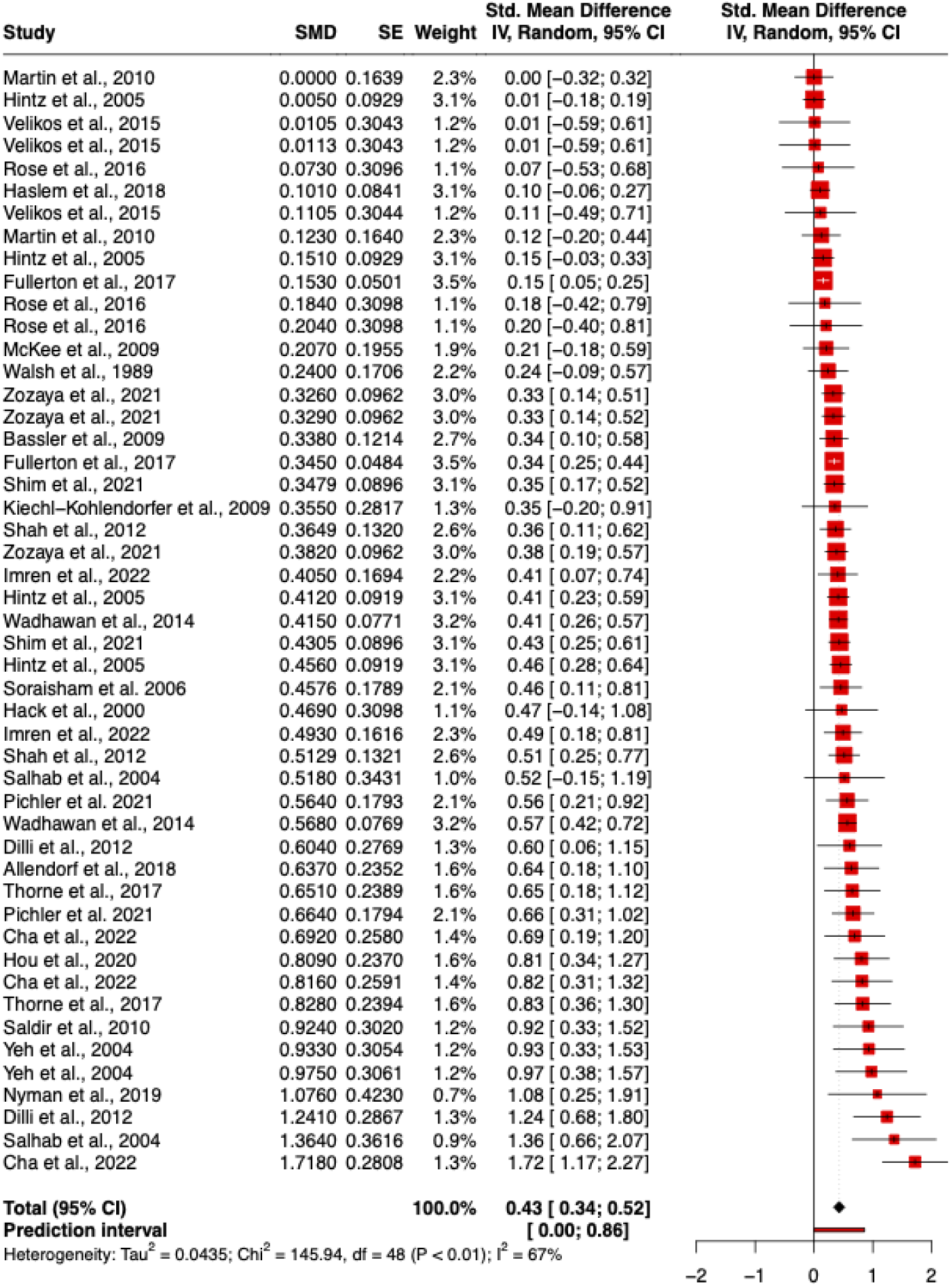
Forest Plot of Pooled Effect Sizes for Bayley Composite Scores (BSID-II and BSID-III). This forest plot presents the pooled effect sizes for Bayley composite scores, combining data from 16 unique studies with Bayley-II scores or a mix of Bayley-II and Bayley-III scores, and 7 unique studies with Bayley-III scores only. The overall pooled effect size is 0.43 (95% CI: 0.34 to 0.52), indicating a moderate effect where individuals with NEC have lower Bayley scores compared to those without NEC. The I² statistic of 67% suggests moderate heterogeneity among studies, and Cochran’s Q test (Q = 145.94, p < 0.001) confirms significant variability.

**Figure 3.**
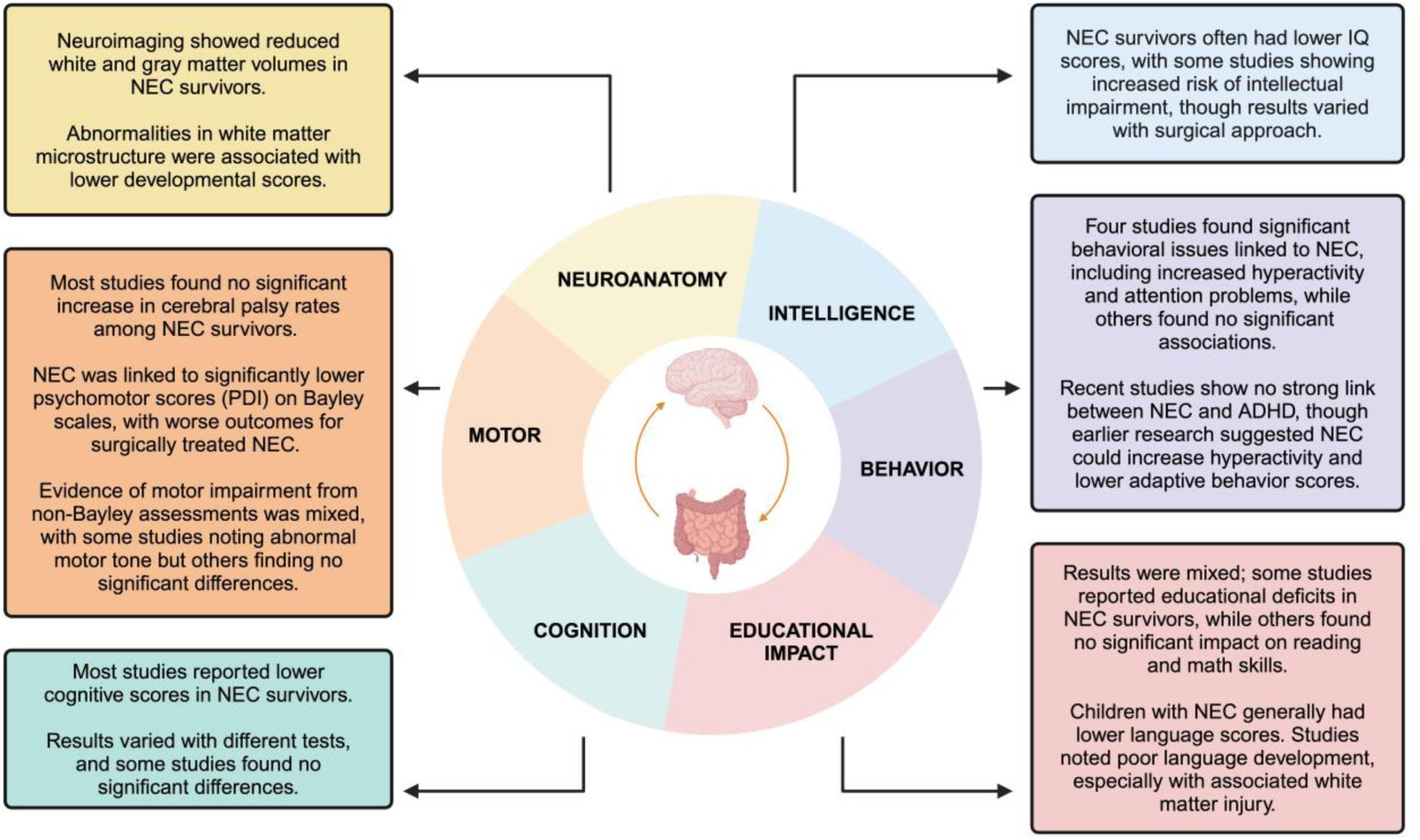
Overview of NEC’s impact on various developmental domains. This graphic provides an at-a-glance view of how NEC influences overall development including general trends and mixed results across different studies. Key sections include motor outcomes, cognitive performance, educational achievement, and neuroanatomical changes.

**Table 2.** Summary of effect size based on gestational age and testing domain.

| Bayley Score/Subscore | Effect size (CI) | I<sup>2</sup> (%) | Cochran's Q (*p*-value) |
| --- | --- | --- | --- |
| Bayley II | 0.44 (0.32-0.55) | 71 | 93.03 (< 0.001) |
| Bayley III | 0.46 (0.27-0.64) | 57 | 37.35 (0.002) |
| Composite (II and III) | 0.43 (0.34-0.52) | 67 | 145.94 (< 0.001) |
| *Gestational age\** |  |  |  |
| Extremely preterm | 0.36 (0.24-0.44) | 66 | 80.42 (< 0.001) |
| Very preterm | 0.56 (0.36-0.76) | 63 | 18.71 (0.03) |
| *Age at Bayley testing* |  |  |  |
| < 18 months | 0.56 (0.21-0.90) | 73 | 36.38 (< 0.001) |
| > 18 months | 0.39 (0.31-0.48) | 65 | 105.47 (< 0.001) |
| Motor | 0.58 (0.33-0.84) | 81 | 72.43 (< 0.001) |
| Mental | 0.40 (0.29-0.50) | 57 | 44.59 (< 0.001) |
| Cognitive | 0.36 (0.21-0.50) | 2 | 5.10 (0.40) |
| Language | 0.34 (0.09-0.59) | 0 | 2.70 (0.44) |

\*Extremely preterm: < 28 weeks; Very preterm: 28-31 weeks

## DISCUSSION

Our systematic review and meta-analysis provide a comprehensive evaluation of the neurodevelopmental outcomes associated with necrotizing enterocolitis in infants. The synthesis of current studies reveals consistent associations between NEC and adverse neurodevelopmental trajectories across motor, cognitive, behavioral, and neuroanatomical domains, underscoring the long-term impact of NEC on survivors.

The analysis of motor outcomes indicates that NEC survivors generally exhibit lower psychomotor scores compared to their peers without NEC. Studies included in this review consistently found that infants with NEC, particularly those requiring surgical intervention, are at higher risk for motor impairments. For instance, Wadhawan et al. reported increased odds of neurodevelopmental impairment in surgically treated NEC cases,^34^ and Hintz et al. found that surgical NEC was associated with significantly lower Psychomotor Development Index (PDI) scores on the Bayley Scales of Infant Development (BSID).^36^ Additionally, Allendorf et al. and Martin et al. reported significantly lower PDI scores among NEC survivors, indicating persistent motor deficits such as impaired gross motor skills, delays in reaching developmental milestones, and decreased motor coordination.^31,35^ These results highlight the importance of monitoring and supporting motor development in NEC survivors through early intervention programs. Evidence suggests that physical therapy (PT) and occupational therapy (OT) can improve motor outcomes in preterm infants, including those with a history of NEC, though specific studies on NEC survivors are limited.^71,72^ PT has been shown to enhance gross motor skills and overall development in preterm infants, with early interventions leading to positive outcomes.^73^ Combined interventions involving both PT and OT have demonstrated improvements in motor and cognitive development.^74^ While these findings are promising, further research targeting NEC survivors is needed to optimize early intervention strategies for this population.

Our analyses additionally identified a consistent trend of lower cognitive scores in NEC survivors, with several studies reporting significantly lower IQ scores and increased risk of intellectual impairment. Humberg et al. found a threefold increased risk of subnormal IQ scores in NEC survivors,^30^ a finding echoed by Waugh et al., who reported an increased odds ratio of NEC in infants with intellectual impairment.^33^ Stevenson et al. also found significantly lower cognitive scores in NEC survivors compared to their siblings.^69^ The cognitive challenges faced by NEC survivors are likely due to underlying brain injuries associated with NEC, as suggested by neuroimaging studies revealing structural abnormalities in key brain areas such as white matter, particularly the splenium of the corpus callosum, and diminished gray matter volume.^44,75^

The behavioral outcomes associated with NEC present a more complex and mixed picture. While some studies, such as those by Astbury et al. and Taylor et al.,^49,61^ indicate a significant association between NEC and adverse behavioral outcomes like attention deficit disorder and hyperactivity, others, like Downey et al. and Pike et al.,^29,62^ do not find significant relationships. Astbury et al. found that children with ADD were significantly more likely to have a history of NEC.^61^ Taylor et al. reported increased parent-reported hyperactivity among school-aged children with a history of NEC.^49^ Neuroplasticity, the late growth of the frontal lobe, and individual differences in resilience and recovery offer plausible explanations for these varied outcomes. For instance, the growth and maturation of the frontal lobe, the area of the brain most responsible for executive functions, decision-making, and higher-order cognitive processes, continues well into early childhood. This prolonged developmental trajectory may provide some compensatory reserve, potentially mitigating the long-term impact of early brain injuries associated with NEC.^76^ Studies have demonstrated that neural plasticity and brain maturation processes can minimize the effects of early developmental challenges. In other words, while the frontal lobe, responsible for executive function and behavioral regulation, undergoes significant development and myelination during the early years of life, its extended period of growth and maturation may contribute to functional reserve and compensate for earlier insults.

Neuroimaging findings provide insights into the structural brain abnormalities associated with NEC. Several studies included in our review reported significant reductions in gray and white matter volumes in NEC survivors.^16-19^ These structural differences were associated with lower neurodevelopmental scores, highlighting the potential impact of NEC on brain development. Notably, studies by Stevenson et al. demonstrated significant differences in the microstructure of white matter, particularly in the corpus callosum, which were correlated with lower developmental scores.^69^ Additionally, studies utilizing MRI and EEG, such as those by Chau et al., reported significant abnormalities in brain structure and function in NEC survivors, further emphasizing the need for neuroprotective strategies in managing NEC to mitigate its impact on brain development.^75^

Our meta-analysis revealed a moderate effect size for lower Bayley scores in NEC survivors, indicating consistent developmental delays in this population. The significant heterogeneity among the studies underscores the variability in study designs, populations, and assessment tools, highlighting the need for standardized methodologies in future research. The pooled effect size suggests that NEC has a substantial impact on neurodevelopmental outcomes, necessitating routine developmental screening and early intervention for NEC survivors.

The cascade of events occurring during necrotizing enterocolitis significantly impacts neurodevelopment through a series of interconnected pathophysiological processes. Initially, NEC often leads to hypoxia-ischemia, whereby reduced blood flow results in energy failure and cell death. This is particularly detrimental to oligodendrocytes, the cells responsible for myelination in the central nervous system. Oligodendrocyte damage impairs myelin formation and integrity, which is crucial for efficient neuronal communication and cognitive function.^77^ Reperfusion injury, which occurs upon the restoration of blood flow, exacerbates the damage by increasing oxidative stress and inflammation. The reintroduction of oxygen and nutrients leads to the generation of reactive oxygen species (ROS) and the activation of inflammatory pathways, further compromising neural tissue and exacerbating neuronal injury.^78^ Additionally, bacterial translocation from the gut into the bloodstream can introduce endotoxins and pathogens into the systemic circulation, intensifying the inflammatory response. This systemic inflammation, characterized by elevated levels of pro-inflammatory cytokines such as IL-6, TNF-α, and IL-1β, can disrupt neurodevelopmental processes by promoting neuroinflammation and damaging brain structures.^16^ The sustained inflammatory response in the neonatal period, combined with the critical window of brain growth and differentiation, may lead to alterations in neural circuits that underlie cognitive and behavioral functions. Collectively, these mechanisms interfere with normal brain development and maturation, contributing to the motor, cognitive, and behavioral deficits observed in NEC survivors. The intricate interplay of these factors underscores the complexity of NEC’s impact on neurodevelopment and highlights the need for targeted interventions to mitigate these long-term consequences.

Overall, the integration of results from both the systematic review and meta-analysis underscores the multifaceted impact of NEC on neurodevelopmental outcomes. The consistent findings across motor, cognitive, behavioral, and neuroanatomical domains highlight the critical need for comprehensive follow-up care for NEC survivors. Early identification of developmental delays through routine screening and targeted interventions can help optimize outcomes for these high-risk infants. However, it is essential to recognize that individual differences in genetic factors, early-life experiences, and environmental supports also contribute to variability in developmental outcomes. Resilience and recovery are influenced by a range of factors, including genetic predisposition, the quality of postnatal care, and the presence of supportive family and environmental factors. Additionally, the presence of neuroprotective factors, such as reduced inflammatory responses or enhanced synaptic plasticity, could contribute to more favorable outcomes in some NEC survivors. The variability in individual responses to NEC-related challenges underscores the importance of personalized approaches to monitoring and intervention.

Future research should elucidate the molecular and cellular mechanisms underlying neurodevelopmental impairments associated with NEC, with a focus on co-morbid conditions and neuroprotective strategies. Long-term, large-scale studies utilizing standardized assessment tools are crucial to fully understand the spectrum of neurodevelopmental outcomes in NEC survivors and to develop effective interventions. This comprehensive meta-analysis and systematic review is among the few to provide an in-depth examination of these issues. Future studies should also investigate whether other types of systemic inflammation produce similar biological and behavioral phenotypes.

In conclusion, NEC is linked to significant neurodevelopmental challenges, highlighting the need for early and sustained support for affected infants. Our findings underscore the importance of implementing thorough care strategies, including regular developmental assessments and customized interventions, to improve long-term outcomes for NEC survivors.

## Data Availability

All data in this systematic review and meta-analysis are from published manuscripts.

## ACKNOWLEDGEMENTS

We would like to acknowledge Scott Appel, MS, and Ashika Mani, MS, from the Biostatistics Analysis Center, University of Pennsylvania, Philadelphia, PA, USA, for their invaluable assistance with the meta-analysis conducted in this study.

