## Supplementary Material for "Association between Neonatal Necrotizing Enterocolitis and Neurodevelopmental Outcomes: A Comprehensive Systematic Review and Meta-Analysis"

**Search Strategies**
**Database: Embase (Elsevier) 1947 - present**
Date searched: January 20, 2022
Results: 6303

| Line | Query |
|---|---|
| 1 | 'newborn'/exp |
| 2 | 'prematurity'/exp |
| 3 | 'newborn screening'/exp |
| 4 | newborn*:ti,ab OR neonat*:ti,ab OR 'premature birth':ti,ab OR infant*:ti,ab OR prematur*:ti,ab OR perinat*:ti,ab |
| 5 | #1 OR #2 OR #3 OR #4 |
| 6 | 'necrotizing enterocolitis'/exp |
| 7 | 'sepsis'/exp |
| 8 | 'systemic inflammation'/exp |
| 9 | 'bloodstream infection'/exp |
| 10 | 'Necrotizing enterocolitis':ti,ab OR 'necrotising enterocolitis':ti,ab OR Sepsis:ti,ab OR 'septic shock':ti,ab OR 'septicemia':ti,ab OR Bacteremia:ti,ab OR 'blood stream infection*':ti,ab OR 'bloodstream infection*':ti,ab OR 'Systemic inflammation':ti,ab |
| 11 | #6 OR #7 OR #8 OR #9 OR #10 |
| 12 | 'cognition'/exp |
| 13 | 'cognitive defect'/exp |
| 14 | 'sleep disorder'/exp |
| 15 | 'behavior disorder'/exp |
| 16 | 'autism'/exp |
| 17 | 'anxiety'/exp |
| 18 | 'anxiety disorder'/exp |
| 19 | 'irritability'/exp |
| 20 | 'colic'/exp |
| 21 | 'memory'/exp |
| 22 | 'memory disorder'/exp |
| 23 | 'emotion'/exp |
| 24 | 'affect'/exp |
| 25 | 'mood disorder'/exp |
| 26 | 'temperament'/exp |
| 27 | 'emotion regulation'/exp |
| 28 | 'executive function'/exp |
| 29 | 'cognitive defect'/exp |
| 30 | 'mental health'/exp |

31 'intellectual impairment'/exp

32 'intelligence test'/exp

33 'psychological adjustment'/exp

34 'postnatal development'/exp

35 'developmental disorder'/exp

36 Cognition:ti,ab OR cognitive:ti,ab OR Milestone*:ti,ab OR neurodevelopment*:ti,ab OR Dysregulation:ti,ab OR 'attention deficit':ti,ab OR ADHD:ti,ab OR Autis*:ti,ab OR Anxiety:ti,ab OR Irritability:ti,ab OR irritable:ti,ab OR Colic:ti,ab OR Bayley:ti,ab OR 'bayley s':ti,ab OR Memory:ti,ab OR Emotion*:ti,ab OR Mood*:ti,ab OR 'affective disorder*':ti,ab OR 'affective disturbance*':ti,ab OR Temperament:ti,ab OR 'Emotional Regulation':ti,ab OR 'Self regulation':ti,ab OR 'Executive function*':ti,ab OR 'executive control':ti,ab OR 'Mental health':ti,ab OR 'mental defect':ti,ab OR 'Intellectual disabilit*':ti,ab OR 'intellectual dysfunction':ti,ab OR 'intellectual impairment*':ti,ab OR 'Intelligence quotient':ti,ab OR 'ages & stages':ti,ab OR 'ages and stages':ti,ab OR ASQ:ti,ab OR DAS:ti,ab OR 'Wechsler Primary Preschool Scale':ti,ab OR WPPSI:ti,ab OR 'Abnormal Involuntary Movement Scale':ti,ab OR 'AIMS score':ti,ab OR 'Adaptive behavior':ti,ab OR 'Developmental functioning':ti,ab OR 'Developmental outcomes':ti,ab OR 'developmental delay*':ti,ab OR 'developmental disorder*':ti,ab OR 'developmental disabilit*':ti,ab

37 #12 OR #13 OR #14 OR #15 OR #16 OR #17 OR #18 OR #19 OR #20 OR #21 OR #22 OR #23 OR #24 OR #25 OR #26 OR #27 OR #28 OR #29 OR #30 OR #31 OR #32 OR #33 OR #34 OR #35 OR #36

38 #5 AND #11 AND #37

**Database: Cochrane Library (Wiley): Cochrane Database of Systematic Reviews 1996 - present, Cochrane Central Register of Controlled Trials 1998 – present**
Date searched: January 20, 2022
Results: 368

| Line | Query |
|---|---|
| 1 | [mh "Infant, Newborn"] |
| 2 | [mh "Premature Birth"] |
| 3 | newborn*:ti,ab OR neonat*:ti,ab OR "premature birth":ti,ab OR infant*:ti,ab OR prematur*:ti,ab OR perinat*:ti,ab |
| 4 | #1 OR #2 OR #3 |
| 5 | [mh "Enterocolitis, Necrotizing"] |
| 6 | [mh Sepsis] |
| 7 | [mh "Shock, Septic"] |
| 8 | [mh Bacteremia] |
| 9 | Necrotizing enterocolitis:ti,ab OR "necrotising enterocolitis":ti,ab OR Sepsis:ti,ab OR "septic shock":ti,ab OR septicemia:ti,ab OR Bacteremia:ti,ab OR ("blood stream" NEXT infection*):ti,ab OR ("bloodstream" NEXT infection*):ti,ab OR "Systemic inflammation":ti,ab |
| 10 | #5 OR #6 OR #7 OR #8 OR #9 |
| 11 | [mh Cognition] |
| 12 | [mh "Cognition Disorders"] |
| 13 | [mh "Chronobiology Disorders"] |
| 14 | [mh "Attention Deficit and Disruptive Behavior Disorders"] |
| 15 | [mh "Autism Spectrum Disorder"] |
| 16 | [mh Anxiety] |
| 17 | [mh "Irritable Mood"] |
| 18 | [mh Colic] |
| 19 | [mh Memory] |
| 20 | [mh Emotions] |
| 21 | [mh Affect] |
| 22 | [mh Temperament] |
| 23 | [mh "Emotional Regulation"] |
| 24 | [mh "Executive Function"] |
| 25 | [mh "Cognitive Dysfunction"] |
| 26 | [mh "Mental Health"] |
| 27 | [mh "Intellectual Disability"] |
| 28 | [mh "Intelligence Tests"] |
| 29 | [mh "Wechsler Scales"] |
| 30 | [mh "Adaptation, Psychological"] |
| 31 | [mh "Child Development"] |

32 [mh "Neurodevelopmental Disorders"]

33 Cognition:ti,ab OR cognitive:ti,ab OR Milestone*:ti,ab OR neurodevelopment*:ti,ab OR Dysregulation:ti,ab OR "attention deficit":ti,ab OR ADHD:ti,ab OR Autis*:ti,ab OR Anxiety:ti,ab OR Irritability:ti,ab OR irritable:ti,ab OR Colic:ti,ab OR Bayley:ti,ab OR "bayley s":ti,ab OR Memory:ti,ab OR Emotion*:ti,ab OR Mood*:ti,ab OR ("affective" NEXT disorder*):ti,ab OR ("affective" NEXT disturbance*):ti,ab OR Temperament:ti,ab OR "Emotional Regulation":ti,ab OR "Self regulation":ti,ab OR ("Executive" NEXT function*):ti,ab OR "executive control":ti,ab OR "Mental health":ti,ab OR "mental defect":ti,ab OR ("Intellectual" NEXT disabilit*):ti,ab OR "intellectual dysfunction":ti,ab OR ("intellectual" NEXT impairment*):ti,ab OR "Intelligence quotient":ti,ab OR "ages & stages":ti,ab OR "ages and stages":ti,ab OR ASQ:ti,ab OR DAS:ti,ab OR "Wechsler Primary Preschool Scale":ti,ab OR WPPSI:ti,ab OR "Abnormal Involuntary Movement Scale":ti,ab OR "AIMS score":ti,ab OR "Adaptive behavior":ti,ab OR "Developmental functioning":ti,ab OR "Developmental outcomes":ti,ab OR ("developmental" NEXT delay*):ti,ab OR ("developmental" NEXT disorder*):ti,ab OR ("developmental" NEXT disabilit*):ti,ab

34 #11 OR #12 OR #13 OR #14 OR #15 OR #16 OR #17 OR #18 OR #19 OR #20 OR #21 OR #22 OR #23 OR #24 OR #25 OR #26 OR #27 OR #28 OR #29 OR #30 OR #31 OR #32 OR #33

35 #4 AND #10 AND #34

**Database: CINAHL (EBSCO) 1937 – present**
Date searched: January 20, 2022
Results: 860

| Line | Query |
|---|---|
| 1 | MH "infant, newborn+" OR MH "Childbirth, Premature" |
| 2 | (ZG "infant, newborn: birth-1 month") or (ZG "infant: 1-23 months") |
| 3 | (TI newborn* OR neonat* OR "premature birth" OR infant* OR prematur* OR perinat*) OR (AB newborn* OR neonat* OR "premature birth" OR infant* OR prematur* OR perinat*) |
| 4 | S1 OR S2 OR S3 |
| 5 | MH "Enterocolitis, Necrotizing" |
| 6 | MH "Sepsis+" |
| 7 | (TI "Necrotizing enterocolitis" OR "necrotising enterocolitis" OR Sepsis OR "septic shock" OR "septicemia" OR Bacteremia OR "blood stream infection*" OR "bloodstream infection*" OR "Systemic inflammation") OR (AB "Necrotizing enterocolitis" OR "necrotising enterocolitis" OR Sepsis OR "septic shock" OR "septicemia" OR Bacteremia OR "blood stream infection*" OR "bloodstream infection*" OR "Systemic inflammation") |
| 8 | S5 OR S6 OR S7 |
| 9 | MH "Cognition+" |
| 10 | MH "Cognition Disorders+" |
| 11 | MH "Sleep disorders+" OR MH "Chronobiology Disorders+" |
| 12 | MH "Attention Deficit Hyperactivity Disorder" |
| 13 | MH "Autistic Disorder" |
| 14 | MH "Anxiety Disorders+" |
| 15 | MH "Anxiety" |
| 16 | MH "Colic+" |
| 17 | MH "Affect" |
| 18 | MH "Affective Disorders+" |
| 19 | MH "Memory+" |
| 20 | MH "Emotional Regulation" |
| 21 | MH "Emotions+" |
| 22 | MH "Temperament" |
| 23 | MH "Executive Function" |
| 24 | MH "Mental Health" |
| 25 | MH "Intellectual Disability+" |
| 26 | MH "Developmental Disabilities" |
| 27 | MH "Intelligence Tests" |
| 28 | MH "Wechsler Memory Scale-Revised" |
| 29 | MH "Wechsler Adult Intelligence Scale-Revised" |

30 MH "Adaptation, Psychological+"
31 MH "Child Development Disorders+"
32 MH "Child Development"
33 MH "Neurodevelopment"
34 MH "Mental Disorders Diagnosed in Childhood+"
35 (TI Cognition OR cognitive OR Milestone* OR neurodevelopment* OR Dysregulation OR "attention deficit" OR ADHD OR Autis* OR Anxiety OR Irritability OR irritable OR Colic OR Bayley OR "bayley s" OR Memory OR Emotion* OR Mood* OR "affective disorder*" OR "affective disturbance*" OR Temperament OR "Emotional Regulation" OR "Self regulation" OR "Executive function*" OR "executive control" OR "Mental health" OR "mental defect" OR "Intellectual disabilit*" OR "intellectual dysfunction" OR "intellectual impairment*" OR "Intelligence quotient" OR "ages & stages" OR "ages and stages" OR ASQ OR DAS OR "Wechsler Primary Preschool Scale" OR WPPSI OR "Abnormal Involuntary Movement Scale" OR "AIMS score" OR "Adaptive behavior" OR "Developmental functioning" OR "Developmental outcomes" OR "developmental delay*" OR "developmental disorder*" OR "developmental disabilit*") OR (AB Cognition OR cognitive OR Milestone* OR neurodevelopment* OR Dysregulation OR "attention deficit" OR ADHD OR Autis* OR Anxiety OR Irritability OR irritable OR Colic OR Bayley OR "bayley s" OR Memory OR Emotion* OR Mood* OR "affective disorder*" OR "affective disturbance*" OR Temperament OR "Emotional Regulation" OR "Self regulation" OR "Executive function*" OR "executive control" OR "Mental health" OR "mental defect" OR "Intellectual disabilit*" OR "intellectual dysfunction" OR "intellectual impairment*" OR "Intelligence quotient" OR "ages & stages" OR "ages and stages" OR ASQ OR DAS OR "Wechsler Primary Preschool Scale" OR WPPSI OR "Abnormal Involuntary Movement Scale" OR "AIMS score" OR "Adaptive behavior" OR "Developmental functioning" OR "Developmental outcomes" OR "developmental delay*" OR "developmental disorder*" OR "developmental disabilit*")
36 S9 OR S10 OR S11 OR S12 OR S13 OR S14 OR S15 OR S16 OR S17 OR S18 OR S19 OR S20 OR S21 OR S22 OR S23 OR S24 OR S25 OR S26 OR S27 OR S28 OR S29 OR S30 OR S31 OR S32 OR S33 OR S34 OR S35
37 S4 AND S8 AND S36

**Database: PsycInfo (EBSCO) 1800s - present**
Date searched: January 20, 2022
Results: 280

| Line | Query |
| --- | --- |
| 1 | DE "Premature Birth" |
| 2 | ZG "neonatal (birth-1 mo)" OR ZG "infancy (2-23 mo)" |
| 3 | DE "Infant Development" OR DE "Infant Temperament" OR DE "Neonatal Development" |
| 4 | (TI newborn* OR neonat* OR “premature birth” OR infant* OR prematur* OR perinat*) OR (AB newborn* OR neonat* OR “premature birth” OR infant* OR prematur* OR perinat*) |
| 5 | S1 OR S2 OR S3 OR S4 |
| 6 | DE "Bacterial Disorders" OR DE "Bacterial Meningitis" OR DE "Gonorrhea" OR DE "Lyme Disease" OR DE "Tuberculosis" |
| 7 | MA Enterocolitis, Necrotizing OR MA Sepsis OR MA shock, Septic |
| 8 | (TI “Necrotizing enterocolitis” OR "necrotising enterocolitis" OR Sepsis OR “septic shock” OR “septicemia” OR Bacteremia OR “blood stream infection*” OR “bloodstream infection*” OR “Systemic inflammation”) OR (AB “Necrotizing enterocolitis” OR "necrotising enterocolitis" OR Sepsis OR “septic shock” OR “septicemia” OR Bacteremia OR “blood stream infection*” OR “bloodstream infection*” OR “Systemic inflammation”) |
| 9 | S6 OR S7 OR S8 |
| 10 | DE "cognition" |
| 11 | DE "Cognitive Ability" |
| 12 | DE "cognitive impairment" |
| 13 | DE "Neurocognitive Disorders" OR DE "Consciousness Disorders" OR DE "Delirium" OR DE "Dementia" OR DE "Memory Disorders" OR DE "Mild Cognitive Impairment" OR DE "Consciousness Disorders" OR DE "Dementia" OR DE "Dementia with Lewy Bodies" OR DE "Presenile Dementia" OR DE "Pseudodementia" OR DE "Semantic Dementia" OR DE "Senile Dementia" OR DE "Vascular Dementia" OR DE "Memory Disorders" OR DE "Amnesia" |
| 14 | DE "Sleep Wake Disorders" OR DE "Hypersomnia" OR DE "Insomnia" OR DE "Narcolepsy" OR DE "Parasomnias" OR DE "Sleep Apnea" |
| 15 | DE "Attention Deficit Disorder" OR DE "Attention Deficit Disorder with Hyperactivity" |
| 16 | DE "Autism Spectrum Disorders" OR DE "Autistic Traits" |
| 17 | DE "Anxiety Disorders" OR DE "Castration Anxiety" OR DE "Generalized Anxiety Disorder" OR DE "Obsessive Compulsive Disorder" OR DE "Panic Attack" OR DE "Panic Disorder" OR DE "Phobias" OR DE "Separation Anxiety Disorder" OR DE "Trichotillomania" |
| 18 | DE "Anxiety" OR DE "Anxiety Sensitivity" OR DE "Computer Anxiety" OR DE "Death Anxiety" OR DE "Health Anxiety" OR DE "Mathematics Anxiety" OR DE "Performance Anxiety" OR DE "Social Anxiety" OR DE "Speech Anxiety" OR DE "Test Anxiety" |

19 DE "Irritability"

20 DE "Affective Disorders" OR DE "Disruptive Mood Dysregulation Disorder" OR DE "Major Depression" OR DE "Anaclitic Depression" OR DE "Dysthymic Disorder" OR DE "Endogenous Depression" OR DE "Late Life Depression" OR DE "Reactive Depression" OR DE "Recurrent Depression" OR DE "Treatment Resistant Depression" OR DE "Seasonal Affective Disorder"

21 DE "Memory" OR DE "Associative Memory" OR DE "Autobiographical Memory" OR DE "Collective Memory" OR DE "Early Memories" OR DE "Eidetic Imagery" OR DE "Episodic Memory" OR DE "Explicit Memory" OR DE "False Memory" OR DE "Forgetting" OR DE "Implicit Memory" OR DE "Long Term Memory" OR DE "Memory Consolidation" OR DE "Memory Decay" OR DE "Memory Trace" OR DE "Prospective Memory" OR DE "Reminiscence" OR DE "Repressed Memory" OR DE "Retrospective Memory" OR DE "Short Term Memory" OR DE "Spatial Memory" OR DE "Spontaneous Recovery (Learning)" OR DE "Tip of the Tongue Phenomenon" OR DE "Verbal Memory" OR DE "Visual Memory"

22 DE "Emotions" OR DE "Contempt" OR DE "Desire" OR DE "Emotional Content" OR DE "Emotional Disturbances" OR DE "Emotional Health" OR DE "Emotional Processing" OR DE "Emotional Regulation" OR DE "Emotional States" OR DE "Emotional Style" OR DE "Emotional Support" OR DE "Expressed Emotion" OR DE "Forgiveness" OR DE "Negative Emotions" OR DE "Positive Emotions" OR DE "Emotional States" OR DE "Affection" OR DE "Agitation" OR DE "Alienation" OR DE "Ambivalence" OR DE "Anger" OR DE "Anxiety" OR DE "Apathy" OR DE "Aversion" OR DE "Belonging" OR DE "Bereavement" OR DE "Boredom" OR DE "Catastrophizing" OR DE "Contentment" OR DE "Depression (Emotion)" OR DE "Disappointment" OR DE "Disgust" OR DE "Dissatisfaction" OR DE "Distress" OR DE "Doubt" OR DE "Embarrassment" OR DE "Emotional Exhaustion" OR DE "Emotional Trauma" OR DE "Enthusiasm" OR DE "Euphoria" OR DE "Euthymia" OR DE "Fear" OR DE "Frustration" OR DE "Gratitude" OR DE "Greed" OR DE "Grief" OR DE "Guilt" OR DE "Happiness" OR DE "Helplessness" OR DE "Homesickness" OR DE "Hope" OR DE "Hopelessness" OR DE "Jealousy" OR DE "Loneliness" OR DE "Love" OR DE "Mania" OR DE "Mental Confusion" OR DE "Morale" OR DE "Optimism" OR DE "Passion" OR DE "Pessimism" OR DE "Pleasure" OR DE "Pride" OR DE "Psychological Capital" OR DE "Psychological Engagement" OR DE "Regret" OR DE "Restlessness" OR DE "Sadness" OR DE "Shame" OR DE "Solidarity" OR DE "Suffering" OR DE "Suspicion" OR DE "Sympathy" OR DE "Emotional Style" OR DE "Emotional States"

23 DE "Personality" OR DE "Personality Processes" OR DE "Behavioral Disinhibition" OR DE "Boundaries (Psychological)" OR DE "Catharsis" OR DE "Cathexis" OR DE "Defense Mechanisms" OR DE "Externalization" OR DE "Identification" OR DE "Inhibition (Personality)" OR DE "Insight" OR DE "Internalization" OR DE "Introspection" OR DE "Personality Change" OR DE "Personality Traits" OR DE "Adaptability (Personality)" OR DE "Aggressiveness" OR DE "Agreeableness" OR DE "Alexithymia" OR DE "Allocentrism" OR DE "Altruism" OR DE "Androgyny" OR DE "Assertiveness" OR DE "Authenticity" OR DE "Authoritarianism" OR DE "Behavioral Inhibition" OR DE "Callous-Unemotional Traits" OR DE "Catastrophizing" OR DE "Charisma" OR DE "Cognitive Style" OR DE "Conformity (Personality)" OR DE

"Conscientiousness" OR DE "Conservatism" OR DE "Courage" OR DE "Cowardice" OR DE "Creativity" OR DE "Cruelty" OR DE "Curiosity" OR DE "Cynicism" OR DE "Dark Triad" OR DE "Defensiveness" OR DE "Dependency (Personality)" OR DE "Dishonesty" OR DE "Dogmatism" OR DE "Egalitarianism" OR DE "Egocentrism" OR DE "Egotism" OR DE "Emotional Immaturity" OR DE "Emotional Inferiority" OR DE "Emotional Instability" OR DE "Emotional Maturity" OR DE "Emotional Security" OR DE "Emotional Stability" OR DE "Emotional Superiority" OR DE "Emotionality (Personality)" OR DE "Empathy" OR DE "Extraversion" OR DE "Femininity" OR DE "Gregariousness" OR DE "Honesty" OR DE "Humility" OR DE "Hypnotic Susceptibility" OR DE "Independence (Personality)" OR DE "Individuality" OR DE "Initiative" OR DE "Integrity" OR DE "Internal External Locus of Control" OR DE "Intolerance of Uncertainty" OR DE "Introversion" OR DE "Irritability" OR DE "Liberalism" OR DE "Likability" OR DE "Loyalty" OR DE "Machiavellianism" OR DE "Masculinity" OR DE "Misanthropy" OR DE "Moodiness" OR DE "Narcissism" OR DE "Need for Approval" OR DE "Need for Cognition" OR DE "Negativism" OR DE "Nervousness" OR DE "Neuroticism" OR DE "Nonconformity (Personality)" OR DE "Nurturance" OR DE "Obedience" OR DE "Objectivity" OR DE "Omnipotence" OR DE "Openmindedness" OR DE "Openness to Experience" OR DE "Optimism" OR DE "Paranoia" OR DE "Passiveness" OR DE "Perceptiveness (Personality)" OR DE "Perfectionism" OR DE "Persistence" OR DE "Pessimism" OR DE "Playfulness" OR DE "Positivism" OR DE "Psychoticism" OR DE "Rebelliousness" OR DE "Repression Sensitization" OR DE "Resilience (Psychological)" OR DE "Rigidity (Personality)" OR DE "Risk Taking" OR DE "Schizotypy" OR DE "Self-Control" OR DE "Self-Regulation" OR DE "Selfishness" OR DE "Sensation Seeking" OR DE "Sensitivity (Personality)" OR DE "Seriousness" OR DE "Sexuality" OR DE "Sincerity" OR DE "Skepticism" OR DE "Sociability" OR DE "Stoicism" OR DE "Subjectivity" OR DE "Suggestibility" OR DE "Timidity" OR DE "Tolerance" OR DE "Psychoanalytic Personality Factors" OR DE "Conscience" OR DE "Conscious (Personality Factor)" OR DE "Death Instinct" OR DE "Ego" OR DE "Electra Complex" OR DE "Fixation (Psychoanalytic)" OR DE "Id" OR DE "Libido" OR DE "Oedipal Complex" OR DE "Subconscious" OR DE "Superego" OR DE "Transitional Objects" OR DE "Unconscious (Personality Factor)" OR DE "Self-Concept" OR DE "Academic Self Concept" OR DE "Entitlement (Psychological)" OR DE "Self-Confidence" OR DE "Self-Congruence" OR DE "Self-Esteem" OR DE "Self-Forgiveness" OR DE "Self-Regard" OR DE "Self-Worth" OR DE "Sense of Coherence"

24 DE "Executive Function" OR DE "Cognitive Control" OR DE "Set Shifting" OR DE "Task Switching"

25 DE "Mental Health" OR DE "Mental Status"

26 DE "Intellectual Development Disorder" OR DE "Mental Retardation"

27 DE "Intelligence Measures" OR DE "Benton Revised Visual Retention Test" OR DE "Culture Fair Intelligence Test" OR DE "Frostig Developmental Test of Visual Perception" OR DE "Goodenough Harris Draw A Person Test" OR DE "Illinois Test of Psycholinguistic Abilities" OR DE "Kaufman Assessment Battery for Children" OR DE "Kohs Block Design Test" OR DE "Miller Analogies Test" OR DE "Peabody Picture Vocabulary Test" OR DE "Porteus Maze Test" OR DE "Raven Coloured Progressive Matrices" OR DE "Raven Progressive Matrices" OR DE "Slosson Intelligence Test" OR

DE "Stanford Binet Intelligence Scale" OR DE "Wechsler Adult Intelligence Scale" OR DE "Wechsler Bellevue Intelligence Scale" OR DE "Wechsler Intelligence Scale for Children" OR DE "Wechsler Preschool Primary Scale"

28 DE "Adjustment" OR DE "Emotional Adjustment" OR DE "Emotional Control" OR DE "Anger control"

29 DE "Adjustment Disorders"

30 DE "Neurodevelopmental Disorders" OR DE "Attention Deficit Disorder" OR DE "Autism Spectrum Disorders" OR DE "Developmental Disabilities" OR DE "Disruptive Behavior Disorders" OR DE "Emotional and Behavioral Disorders" OR DE "Intellectual Development Disorder" OR DE "Learning Disorders" OR DE "Developmental Disabilities" OR DE "Specific Language Impairment" OR DE "Disruptive Behavior Disorders" OR DE "Conduct Disorder" OR DE "Oppositional Defiant Disorder" OR DE "Learning Disorders" OR DE "Learning Disabilities" OR DE "Reading Disabilities"

31 (TI Cognition OR cognitive OR Milestone* OR neurodevelopment* OR Dysregulation OR "attention deficit" OR ADHD OR Autis* OR Anxiety OR Irritability OR irritable OR Colic OR Bayley OR "bayley s" OR Memory OR Emotion* OR Mood* OR "affective disorder*" OR "affective disturbance*" OR Temperament OR "Emotional Regulation" OR "Self regulation" OR "Executive function*" OR "executive control" OR "Mental health" OR "mental defect" OR "Intellectual disabilit*" OR "intellectual dysfunction" OR "intellectual impairment*" OR "Intelligence quotient" OR "ages & stages" OR "ages and stages" OR ASQ OR DAS OR "Wechsler Primary Preschool Scale" OR WPPSI OR "Abnormal Involuntary Movement Scale" OR "AIMS score" OR "Adaptive behavior" OR "Developmental functioning" OR "Developmental outcomes" OR "developmental delay*" OR "developmental disorder*" OR "developmental disabilit*") OR (AB Cognition OR cognitive OR Milestone* OR neurodevelopment* OR Dysregulation OR "attention deficit" OR ADHD OR Autis* OR Anxiety OR Irritability OR irritable OR Colic OR Bayley OR "bayley s" OR Memory OR Emotion* OR Mood* OR "affective disorder*" OR "affective disturbance*" OR Temperament OR "Emotional Regulation" OR "Self regulation" OR "Executive function*" OR "executive control" OR "Mental health" OR "mental defect" OR "Intellectual disabilit*" OR "intellectual dysfunction" OR "intellectual impairment*" OR "Intelligence quotient" OR "ages & stages" OR "ages and stages" OR ASQ OR DAS OR "Wechsler Primary Preschool Scale" OR WPPSI OR "Abnormal Involuntary Movement Scale" OR "AIMS score" OR "Adaptive behavior" OR "Developmental functioning" OR "Developmental outcomes" OR "developmental delay*" OR "developmental disorder*" OR "developmental disabilit*")

32 S10 OR S11 OR S12 OR S13 OR S14 OR S15 OR S16 OR S17 OR S18 OR S19 OR S20 OR S21 OR S22 OR S23 OR S24 OR S25 OR S26 OR S27 OR S28 OR S29 OR S30 OR S31

33 S5 AND S9 AND S32

**Database: MEDLINE (Pubmed) 1946 - present**
Date searched: January 20, 2022
Results: 1833

| Line | Query |
|---|---|
| 1 | "Infant, Newborn"[Mesh] |
| 2 | Premature Birth[Mesh] |
| 3 | newborn*[tiab] OR neonat*[tiab] OR “premature birth”[tiab] OR infant*[tiab] OR prematur*[tiab] OR perinat*[tiab] |
| 4 | #1 OR #2 OR #3 |
| 5 | "Enterocolitis, Necrotizing"[Mesh] |
| 6 | "Sepsis"[Mesh] |
| 7 | "Shock, Septic"[Mesh] |
| 8 | "Bacteremia"[Mesh] |
| 9 | “Necrotizing enterocolitis”[tiab] OR "necrotising enterocolitis"[tiab] OR Sepsis[tiab] OR “septic shock”[tiab] OR “septicemia”[tiab] OR Bacteremia[tiab] OR “blood stream infection*”[tiab] OR “bloodstream infection*”[tiab] OR “Systemic inflammation”[tiab] |
| 10 | #5 OR #6 OR #7 OR #8 OR #9 |
| 11 | "Cognition"[Mesh] |
| 12 | "Cognition Disorders"[Mesh] |
| 13 | "Chronobiology Disorders"[Mesh] |
| 14 | "Attention Deficit and Disruptive Behavior Disorders"[Mesh] |
| 15 | "Autism Spectrum Disorder"[Mesh] |
| 16 | "Anxiety"[Mesh] |
| 17 | "Irritable Mood"[Mesh] |
| 18 | "Colic"[Mesh] |
| 19 | "Memory"[Mesh] |
| 20 | "Emotions"[Mesh] |
| 21 | "Affect"[Mesh] |
| 22 | "Temperament"[Mesh] |
| 23 | "Emotional Regulation"[Mesh] |
| 24 | "Executive Function"[Mesh] |
| 25 | "Cognitive Dysfunction"[Mesh] |
| 26 | "Mental Health"[Mesh] |
| 27 | "Intellectual Disability"[Mesh] |
| 28 | "Intelligence Tests"[Mesh] |
| 29 | "Wechsler Scales"[Mesh] |
| 30 | "Adaptation, Psychological"[Mesh] |
| 31 | “Child Development”[MeSH] |
| 32 | "Neurodevelopmental Disorders"[Mesh] |

| | |
|---|---|
| 33 | Cognition[tiab] OR cognitive[tiab] OR Milestone*[tiab] OR neurodevelopment*[tiab] OR Dysregulation[tiab] OR "attention deficit"[tiab] OR ADHD[tiab] OR Autis*[tiab] OR Anxiety[tiab] OR Irritability[tiab] OR irritable[tiab] OR Colic[tiab] OR Bayley[tiab] OR "bayley s"[tiab] OR Memory[tiab] OR Emotion*[tiab] OR Mood*[tiab] OR "affective disorder*"[tiab] OR "affective disturbance*"[tiab] OR Temperament[tiab] OR "Emotional Regulation"[tiab] OR "Self regulation"[tiab] OR "Executive function*"[tiab] OR "executive control"[tiab] OR "Mental health"[tiab] OR "mental defect"[tiab] OR "Intellectual disabilit*"[tiab] OR "intellectual dysfunction"[tiab] OR "intellectual impairment*"[tiab] OR "Intelligence quotient"[tiab] OR "ages & stages"[tiab] OR "ages and stages"[tiab] OR ASQ[tiab] OR DAS[tiab] OR "Wechsler Primary Preschool Scale"[tiab] OR WPPSI[tiab] OR "Abnormal Involuntary Movement Scale"[tiab] OR "AIMS score"[tiab] OR "Adaptive behavior"[tiab] OR "Developmental functioning"[tiab] OR "Developmental outcomes"[tiab] OR "developmental delay*"[tiab] OR "developmental disorder*"[tiab] OR "developmental disabilit*"[tiab] |
| 34 | #11 OR #12 OR #13 OR #14 OR #15 OR #16 OR #17 OR #18 OR #19 OR #20 OR #21 OR #22 OR #23 OR #24 OR #25 OR #26 OR #27 OR #28 OR #29 OR #30 OR #31 OR #32 OR #33 |
| 35 | #4 AND #10 AND #34 |
